# Rapid whole-brain high-resolution myelin water fraction mapping from extremely under-sampled magnetic resonance imaging data using deep neural network

**DOI:** 10.1101/2023.03.07.23286946

**Authors:** Zhaoyuan Gong, Nikkita Khattar, Matthew Kiely, Curtis Triebswetter, Mustapha Bouhrara

## Abstract

Changes in myelination are a cardinal feature of brain development and the pathophysiology of several cerebral diseases, including multiple sclerosis and dementias. Advanced magnetic resonance imaging (MRI) methods have been developed to probe myelin content through the measurement of myelin water fraction (MWF). However, the prolonged data acquisition and post-processing times of current MWF mapping methods pose substantial hurdles to their clinical implementation. Recently, fast steady-state MRI sequences have been implemented to produce high spatial resolution whole-brain MWF mapping within ∼ 20 min. Despite the subsequent significant advances in the inversion algorithm to derive MWF maps from steady-state MRI, the high-dimensional nature of such inversion does not permit further reduction of the acquisition time by data under-sampling. In this work, we present an unprecedented reduction in the computation (∼ 30 s) and the acquisition time (∼ 7 min) required for whole-brain high-resolution MWF mapping through a new Neural Network (NN)-based approach, named: Relaxometry of Extremely Under-SamplEd Data (NN-REUSED). Our analyses demonstrate virtually similar accuracy and precision in derived MWF values using the NN-REUSED approach as compared to results derived from the fully-sampled reference method. The reduction in the acquisition and computation times represents a breakthrough toward clinically practical MWF mapping.

## I. INTRODUCTION

Myelin is paramount for the normal functioning of the central nervous system (CNS), with loss or damage of the myelin sheets leading to various neurological diseases, including multiple sclerosis and dementias^1–7^. As an electrical insulator essential for action potential conduction and trophic support to the neuronal axons of the CNS^8^, myelin is crucial to higher-order integrative functions of the brain^9^. Therefore, probing myelin content and its integrity is critical to understanding cerebral development, maturation, and degeneration^10–12^, as well as enhancing our capacity to identify novel therapeutics for myelin repair^13–15^. For that, magnetic resonance imaging (MRI) of myelin water fraction (MWF), a direct proxy of myelin content^16,17^, has been developed based on the multicomponent nature of water pools within image voxels. These water pools exhibit differential nuclear magnetic properties, including relaxation times. The fast relaxing component corresponds to the water trapped within the myelin sheets, while the moderately and slowly relaxing components are attributed to intra/extra cellular and cerebrospinal fluid (CSF) water compartments, respectively^16,17^. The signal fraction of the fast relaxing component is defined as the MWF, which has been histologically validated as a specific measure of myelin content^3,4^.

The multi-echo spin-echo (MESE) sequence was originally used to measure in vivo MWF by Mackay *et al*. ^16^, where the nuclear magnetization is excited by a 90^*◦*^ radio-frequency pulse followed by a train of 180^*◦*^ refocusing pulses. The signal amplitudes, *S*_*i*_, acquired at the *i*-th echo time, *t*_*i*_, follow the form of a sum of several exponential decays through:

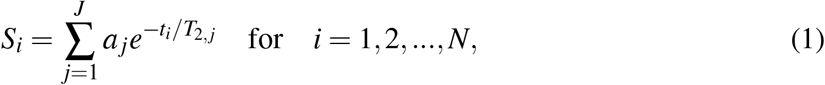

where N is the total number of echoes acquired, and *a* _*j*_ is the *j*-th component of a total of J number of components with distinct transverse relaxation time *T*_2, *j*_ and fraction *a* _*j*_. The decay curve is conventionally inverted using a nonnegative least-squares (NNLS) optimization to obtain the short *T*_2_ relaxation fraction, i.e., MWF. Despite being the gold standard method for MWF mapping, MESE suffers the utmost slow data acquisition and fitting instability. Using fast steady-state MRI sequences, namely the spoiled gradient recalled echo (SPGR) and balanced steady-state free precession (bSSFP) sequences, the multicomponent driven equilibrium steady-state observation of *T*_1_ and *T*_2_ (mcDESPOT)^18–23^ has been introduced for whole-brain high spatial resolution MWF determination within a clinically feasible acquisition time (∼20 min). To improve the accuracy and precision in derived MWF values, the Bayesian Monte Carlo (BMC) analysis of mcDESPOT (BMC-mcDESPOT) was proposed and extensively applied in studies of cerebral maturation and degeneration^12,21,22,24–26^.

Despite these critical advances for clinically feasible and accurate MWF imaging, the acquisition time remains relatively long, particularly infeasible in investigations involving participants with limited cooperability or in studies acquiring other imaging modalities within the same scan session. Further, due to its underlying complex mathematical modeling, BMC-mcDESPOT requires extensive computational power, with several hours needed to generate a single whole-brain MWF map; this limits real-time evaluation. Most importantly, as clarified in (2)-(4) in the Methods section below, estimation of the MWF values using BMC-mcDESPOT involves a high dimensional inversion problem, incorporating several unknown parameters requiring, therefore, several measurements, i.e., several SPGR and bSSFP data at different FAs in this case, for an accurate determination of MWF. Indeed, estimates of MWF from a limited number of SPGR and bSSFP images are expected to be inaccurate and even impossible using BMC-mcDESPOT. In this study, we demonstrate the possibility of extreme data under-sampling in the s-space (referring to the s domain in Laplace transform) for quantitative MRI acquisition using neural network (NN) estimation of MWF. Specifically, we show that derived MWF maps using our NN method from a limited number of SPGR and bSSFP images are virtually identical to those derived from the reference method and a larger number of images.

## II. RELATED WORKS

From partial Fourier techniques based on conjugate symmetry, and compressed sensing algorithms based on data sparsity, to the emerging NN aided imaging reconstruction, data undersampling is mainly explored in the k-space. Building on the same principle, it is expected that NN-based algorithms will also exhibit high performance in parameters estimation from undersampled data in the s-space, allowing accelerated acquisition for quantitative MRI.

Furthermore, emerging evidence^27–36^ demonstrates that artificial NN can also drastically shorten the computational burden in parameters estimation, and was also used to derive MWF maps from a limited number of images. In recent work, Liu *et al*. ^29^ have proposed deep learning NN models for rapid computation of MWF maps from conventional multiple echo-time images^37^. Using advanced NN algorithms, we Khattar *et al*. ^36^ and Piredda *et al*. ^32^ have recently shown the possibility of deriving MWF maps from proton density (PD), longitudinal relaxation (*T*_1_) and transverse relaxation (*T*_2_) parameter maps. Furthermore, Jung *et al*. ^35^ showed that the accuracy in derived MWF values from multiple gradient-echo images is markedly improved using NN as compared to the conventional, widely-used, least-squares fitting algorithms. However, despite these remarkable advances in MWF determination using NN, most of these methods focused on fast computation or providing MWF with a low spatial or temporal resolution. Building on this seminal work, in the following, we will provide details on our NN-based approach for whole-brain high-resolution MWF mapping from steady-state imaging data; namely, SPGR and bSSFP, within drastically reduced acquisition times as compared to the state-of-the-art method, BMC-mcDESPOT.

## III. METHODS

### A. Reference MWF maps generation

#### 1. BMC-mcDESPOT for reference MWF maps

In BMC-mcDESPOT, MWF is estimated from a set of bSSFP and SPGR images. These steady-state images are acquired at several flip angles (FAs) with very short repetition times (TRs), allowing a substantial reduction in the total acquisition time for whole-brain high-resolution imaging as compared to the conventional methods. Using the Bayes theorem and Monte Carlo sampling, the estimate, 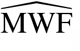, of the parameter, MWF, belonging to the set of unknown parameters ***λ*** is given by:

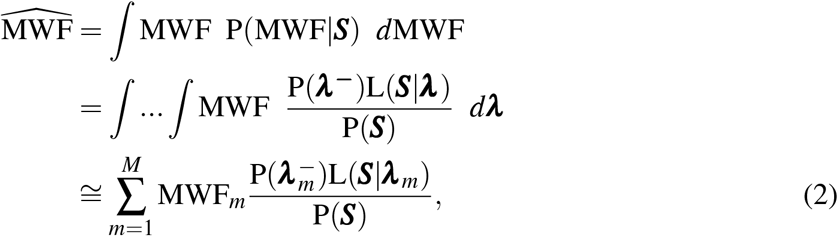

where P(MWF|***S***) is the posterior distribution of MWF, ***S*** = (***S***_SPGR_, ***S***_bSSFP_) is the ensemble of the measured SPGR and bSSFP signal vectors at *K* FAs in the index voxel, *m* denotes one of a total of *M* = 30, 000 random sets of parameter combinations sampled from a grid defining the range of our Monte Carlo integration, P(***S***) = ∫P(***λ*** ^*−*^)L(***S***|***λ***)*d****λ*** is a normalization constant, and ***λ***^*−*^ is equivalent to ***λ*** but excludes the parameter of interest, that is, MWF. Assuming a bicomponent model with a two-relaxation time components system consisting of short, attributed to myelin water, and long, attributed to intra/extra (IE) cellular water, components, ***λ*** = (MWF, *T*_1,MWF_, *T*_1,IE_, *T*_2,MWF_, *T*_2,IE_) is the vector of the unknown parameters, 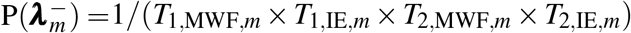 is Jeffrey’s noninformative priors, and L(***S***|***λ*** _*m*_) is the likelihood function of ***S*** given ***λ*** _*m*_ and is given by:

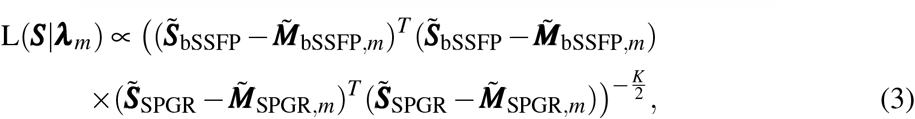

where 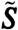 and 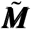 are respectively the experimental and theoretical signals normalized by their respective mean values calculated over *K* SPGR or bSSFP FAs, given by:

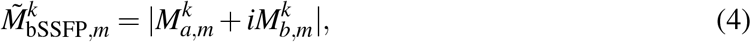

with

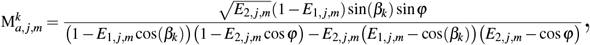

and

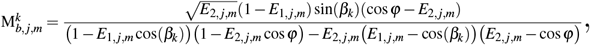

and

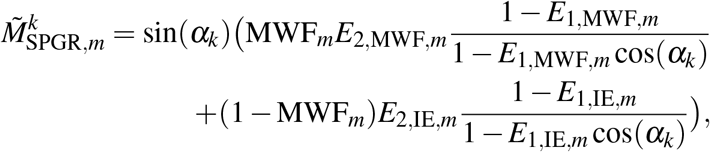

where *ϕ* = 2*π ·* ∆*ω ·* TR_bSSFP_ + *ϑ*, with ∆*ω* the off-resonance frequency of the proton pool and *ϑ* the phase increment of the applied radio-frequency (RF) pulse. *E*_1, *j,m*_ = exp(*−*TR_bSSFP_*/T*_1, *j,m*_) and *E*_2, *j,m*_ = exp(*−*TR_bSSFP_*/T*_2, *j,m*_), where *T*_1, *j*_ and *T*_2, *j*_ are, respectively, the spin-lattice and spin-spin relaxation times of the *j*^th^ component (i.e., MWF or IE). Further details can be found in^20,21,23^.

#### 2. MRI data acquisition

All MRI scans were performed on a 3T whole-body Philips MRI system (Achieva, Best, The Netherlands) using the internal quadrature body coil for transmission and an eight-channel phased-array head coil for signal acquisition. Under the approval of the institutional review board, the following three sequences were acquired in the mcDESPOT protocol to calculate MWF: 3D SPGR images were acquired with FAs of [2 4 6 8 10 12 14 16 18 20]^*◦*^, echo time (TE) of 1.37 ms and repetition time (TR) of 5 ms. 3D bSSFP images were acquired with FAs of [2 4 7 11 16 24 32 40 50 60]^*◦*^, TE of 2.8 ms, and TR of 5.8 ms. The bSSFP images were acquired twice with radiofrequency excitation pulse phase cycling of 0 (bSSFP_0_) and *π* (bSSFP_*π*_) to account for off-resonance effects^19^. All SPGR and bSSFP images were acquired with an acquisition matrix of 150 *×* 130 *×* 94, and a voxel size of 1.6 mm *×*1.6 mm *×* 1.6 mm. To correct for excitation radiofrequency inhomogeneity, *B*_1_, we used the double-angle method (DAM)^38^ by acquiring two fast spin-echo images with FAs of 45^*◦*^ and 90^*◦*^, TE of 102 ms, TR of 3000 ms, and acquisition voxel size of 2.6 mm *×* 2.6 mm *×* 4 mm. All images were acquired with a field of view of 240 mm *×* 208 mm *×* 150 mm. The total acquisition time was ∼21 min, in which SPGR, two bSSFP, and DAM datasets were ∼5 min, ∼12 min, and ∼4 min, respectively.

In this work, we will refer to the mcDESPOT dataset with ten SPGR/bSSFP acquired at different FAs as fully-sampled, while datasets with four FAs as under-sampled. We also consider a dataset with only two FAs as extremely under-sampled.

#### 3. MWF mapping

For each participant, a whole-brain *B*_1_ map was generated from the DAM datasets and extrapolated to the same size as of the bSSFP and SPGR images, and a reference whole-brain MWF map was generated from the fully-sampled SPGR and bSSFP images and the derived *B*_1_ map using BMC-mcDESPOT^21^. In comparison to the NN-REUSED approach described below, we also derived MWF maps using BMC-mcDESPOT from the under-sampled and extremely under-sampled dataset. This is conducted to illustrate the limitation of the BMC-mcDESPOT analysis to generate accurate MWF maps from under-sampled or extremely under-sampled datasets.

#### 4. Image registration

The calculated *B*_1_ and MWF maps were linearly registered to the averaged SPGR image overall FAs for that corresponding participant using the FSL linear image registration tool (FLIRT)^39^. The averaged SPGR image was passed to the FSL brain extraction tool (BET)^40^ to eliminate non-brain structures and to calculate the brain mask. Then it was registered to the Montreal Neurological Institute (MNI) standard space with 1 mm *×* 1 mm *×* 1 mm resolution and matrix size of 182 *×* 218 *×* 182 using the FSL nonlinear image registration tool (FNIRT), and the derived transformation matrix was then applied to *B*_1_ and MWF maps.The same registration process was repeated for all SPGR, bSSFP, and DAM images. Finally, brain tissues were segmented into white matter, gray matter, and cerebrospinal fluid using the FSL automated segmentation tool (FAST)^39^. Voxels belonging to the CSF compartment were excluded from the subsequent model training. Indeed, for the training data, this segmentation to exclude CSF was necessary as we observed that contamination from CSF hinders the convergence of models.

### B. NN Relaxometry of Extremely Under-SamplEd Data (NN-REUSED) implementation

#### 1. The NN-REUSED architecture

Our NN Relaxometry of Extremely Under-SamplEd Data (NN-REUSED) approach is based on the ResNet-like^41^ fully connected networks. Building blocks of the deep NN-REUSED are illustrated in Fig.1 (A). Incoming features (In_features) were passed through three fully connected dense layers with BatchNorm^42^ and LeakyReLu activation in between before being added into the output features (Out_features) if the In_features = Out_features. Otherwise, the In_features was passed through a fully connected layer to change the size of Out_features and then added to the output features. The addition or skip connection here behaves like an information highway and helps with gradient flow, enabling stable training of very deep networks^41^.

**FIG. 1.**
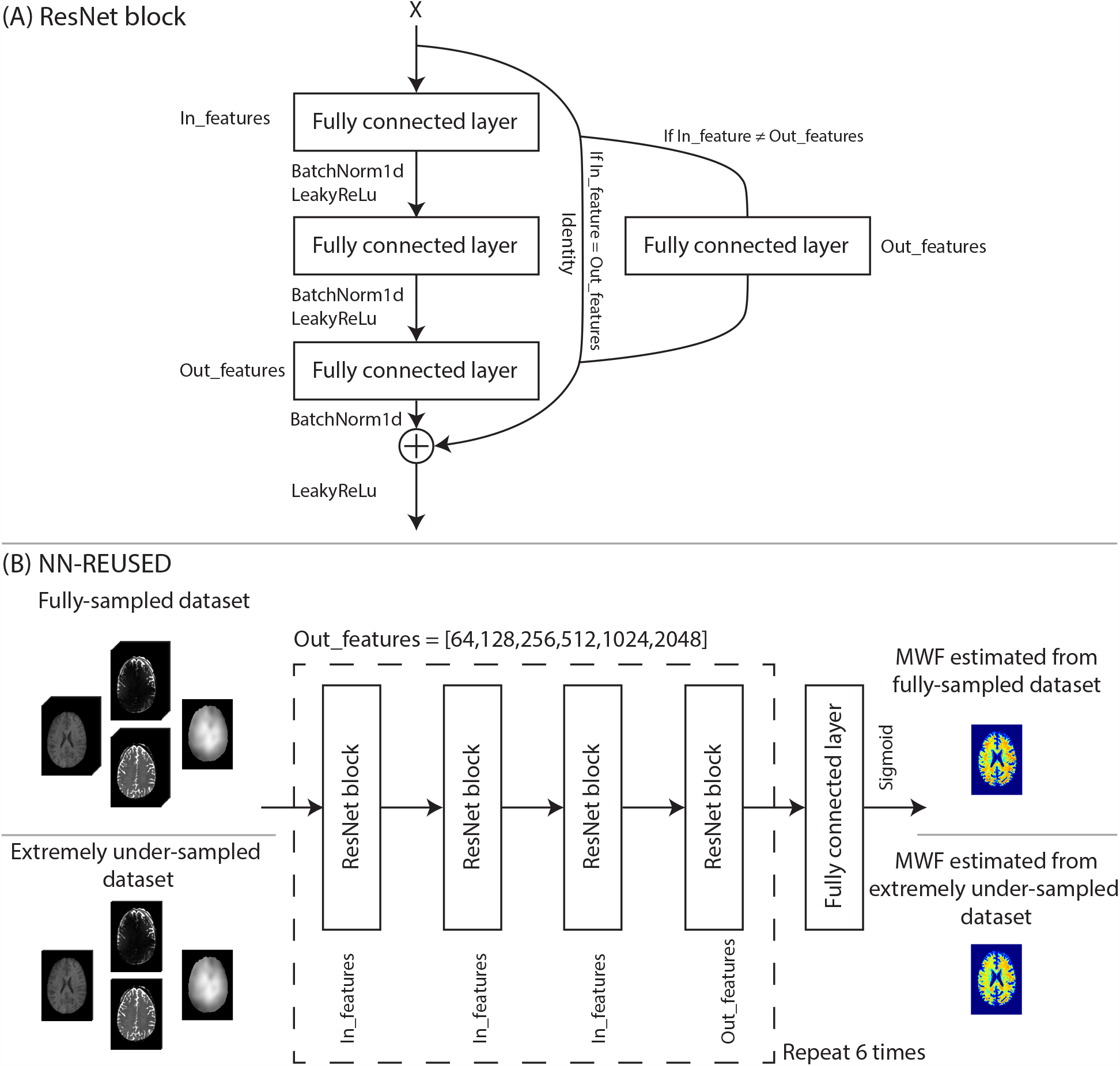
Schematic sketch of (A) the fully connected ResNet building block, (B) the NN-REUSED architecture and the illustration of using either fully-sampled datasets or extremely under-sampled datasets to generate MWF maps.

For the fully-sampled model, the input features for NN-REUSED are the intensity values of the SPGR and bSSFP voxels obtained from 30 SPGR and bSSFP images (i.e., 10 SPGR images, 10 bSSFP_0_ images, and 10 bSSFP_*π*_ images) in addition to the *B*_1_ map. For the under-sampled model, only 12 SPGR and bSSFP images (i.e., 4 SPGR images, 4 bSSFP_0_ images, and 4 bSSFP_*π*_ images) in addition to the *B*_1_ map were used as input features. For the extremely under-sampled model, only 6 SPGR and bSSFP images (i.e., 2 SPGR images, 2 bSSFP_0_ images, and 2 bSSFP_*π*_ images) in addition to the *B*_1_ map were used as input features. For all models (Fig.1 (B)), between the input and output layers, there are 24 ResNet blocks, with the output features changing size every four blocks to 64, 128, 256, 512, 1024, and 2048, respectively. The model optimizer is Adam^43^, and the loss function is L1 loss. The learning rate is initially set to 0.0001 and decreased ten folds after each epoch along with gradient clipping setting maximum gradient values to be 0.0001.

These measures greatly stabilized the networks, and models were trained for 200 epochs. All codes were implemented in PyTorch^44^ version 1.9.0, and all training and testing were performed on a Dell precision workstation equipped with Intel^®^ Xeon^®^ gold 6230 CPU @ 2.10 GHz and NVIDIA Quadro RTX™ 5000 GPU. Data and codes are available upon request from the first and last authors.

#### 2. Model training

MRI datasets of 37 cognitive normal subjects ranging from 22 to 94 years (mean = 56.9, SD = 23.7) were randomly chosen from the Baltimore Longitudinal Study of Aging (BLSA). After image registration, all calculated maps and raw images were multiplied by the WM and GM masks of each participant to retain WM and GM tissues only and exclude CSF. All maps and raw images were vectorized to create the input features. All reference MWF maps were calculated using the BMC-mcDESPOT method from the fully-sampled datasets^21^. There are 78 million final available pairs of features and labels, while random split ratio between training and validation is 9:1.

#### 3. Model testing

The testing datasets constitute 39 cognitive normal subjects ranging from 24 to 94 years old (mean = 60.7, SD = 22.7). To test the accuracy and utility of the NN-REUSED approach in estimating MWF maps from data obtained from participants with neurodegeneration, the model testing also included three patients with Alzheimer’s disease (AD). We note that our NN-REUSED models were solely trained on cognitively normal participants. After image registration, the calculated maps and raw images were masked out of CSF and vectorized to create the input features. Finally, the calculated MWF values using the NN-REUSED models were reshaped to the original three dimensional size providing corresponding whole-brain MWF maps. Exemplary results from one 75-year-old male AD patient and one healthy 33-year-old female subject are shown in Fig.2, 3, 4.

**FIG. 2.**
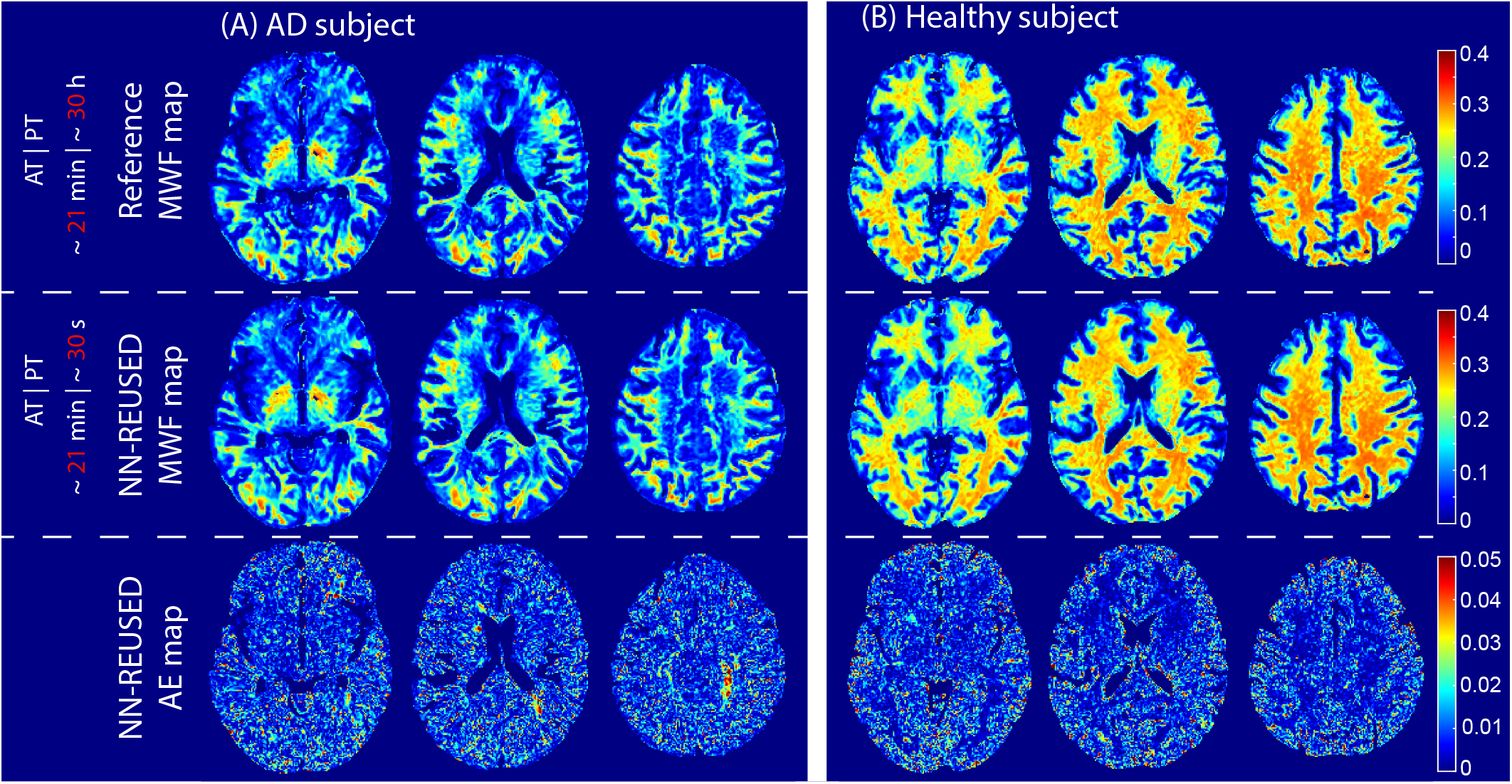
Examples of MWF maps derived using BMC-mcDESPOT (first row, Reference map) or NN-REUSED (second row) from the fully-sampled datasets of an AD patient (A) and a healthy subject (B). Corresponding absolute error (AE) maps calculated as the absolute difference between the MWF map derived using BMC-mcDESPOT and the MWF map derived using NN-REUSED are also displayed (bottom row). Results are shown for three representative axial slices. The acquisition time (AT) and processing time (PT) are labeled in red to show the time accelerations.

**FIG. 3.**
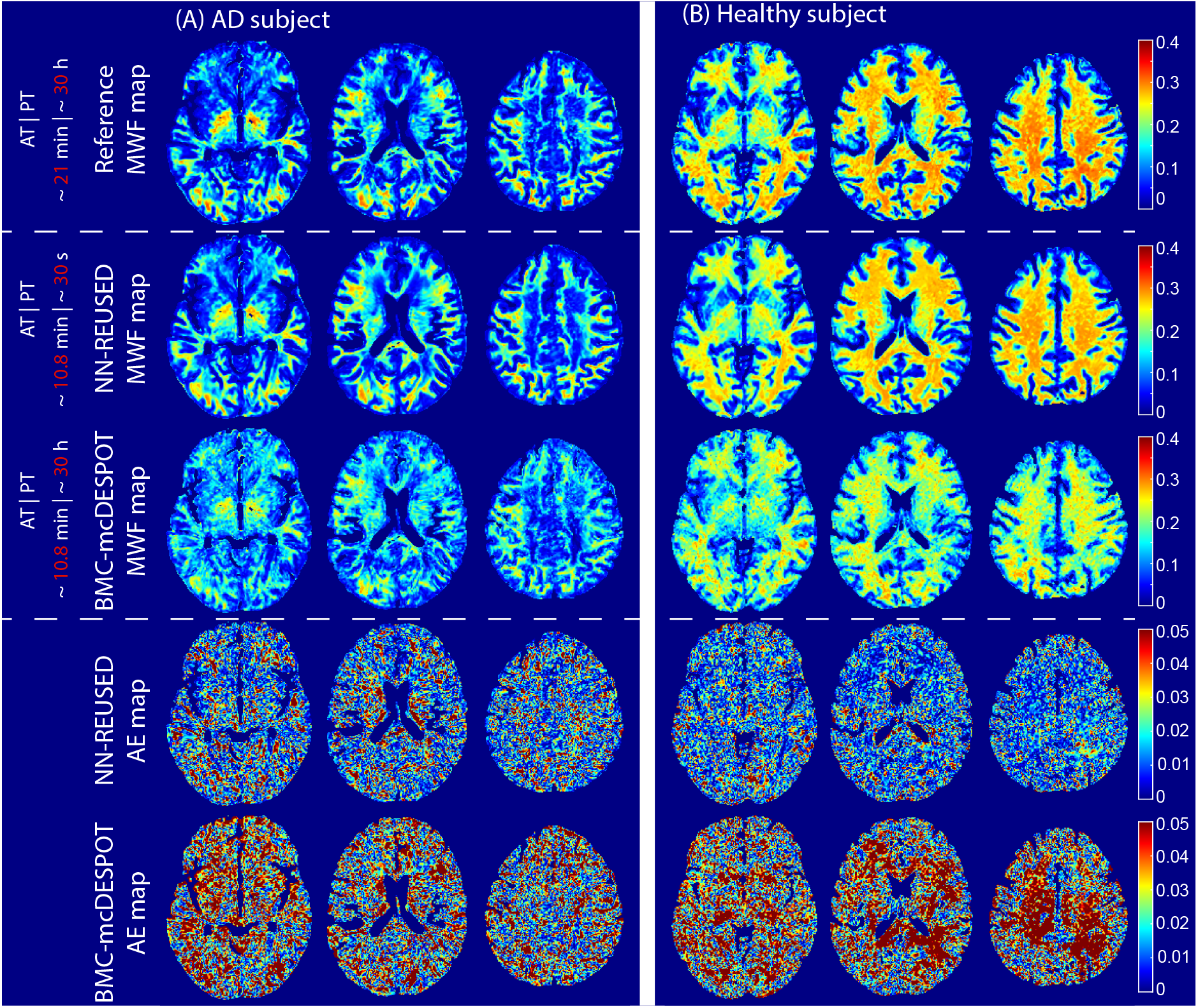
Examples of derived MWF maps using the reference method (i.e., BMC-mcDESPOT from the fully-sampled datasets, first row), NN-REUSED (second row) or BMC-mcDESPOT (third row) from under-sampled datasets of an AD patient (A) and a healthy subject (B). Corresponding absolute error (AE) maps calculated as the absolute difference between the MWF map derived using the references method and the MWF map derived using either NN-REUSED or BMC-mcDESPOT from under-sampled datasets are also displayed (bottom rows). Results are shown for three representative axial slices. The acquisition time (AT) and processing time (PT) are labeled in red to show the time accelerations.

**FIG. 4.**
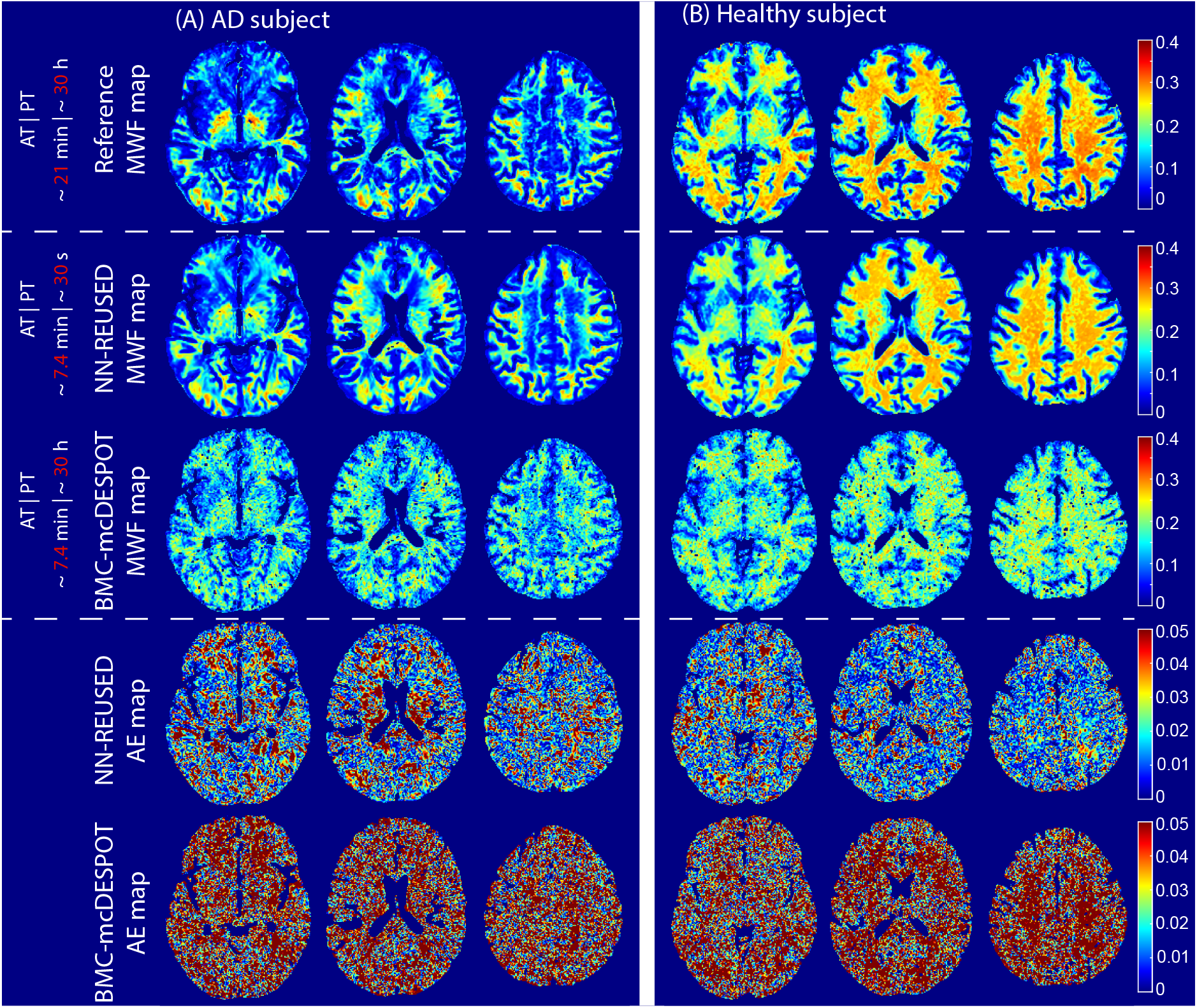
Examples of derived MWF maps using the reference method (i.e., BMC-mcDESPOT from the fully-sampled datasets, first row), NN-REUSED (second row) or BMC-mcDESPOT (third row) from extremely under-sampled datasets of an AD patient (A) and a healthy subject (B). Corresponding absolute error (AE) maps calculated as the absolute difference between the MWF map derived using the references method and the MWF map derived using either NN-REUSED or BMC-mcDESPOT from extremely under-sampled datasets are also displayed (bottom rows). Results are shown for three representative axial slices. The acquisition time (AT) and processing time (PT) are labeled in red to show the time accelerations.

### C. BMC-mcDESPOT and NN-REUSED methods comparison

#### 1. Absolute error maps

The absolute error (AE) maps were calculated by taking the absolute difference between the estimated MWF and reference MWF maps. A restricted color scale has been used to highlight the superior performance of NN-REUSED over BMC-mcDESPOT for under-sampled and extremely under-sampled datasets.

#### 2. Bland-Altman plot and linear correlation plot of regions-of-interest

Six WM and GM structures were chosen as regions of interest (ROIs) from the MNI atlas provided in FSL. These regions were defined from the Johns Hopkins University (JHU) ICM-DTI-81 atlas. For each participant, all ROIs were eroded to reduce partial volume effects and the mean MWF values from BMC-mcDESPOT or NN-REUSED were calculated. Bland-Altman plots and linear correlation plots of 12 ROIs for fully-sampled, under-sampled, or extremely under-sampled datasets were generated using MATLAB.

### D. Structure similarity analysis

Structural similarity index measure (SSIM) between MWF maps generated from the NN-REUSED and reference maps were calculated for each testing dataset using the MATLAB function ssim. Results are shown in Fig.5, 6. The mean and standard deviation SSIM values calculated across all testing participants were also generated. For each testing dataset, results are shown for the fully-sampled, under-sampled and extremely under-sampled datasets.

**FIG. 5.**
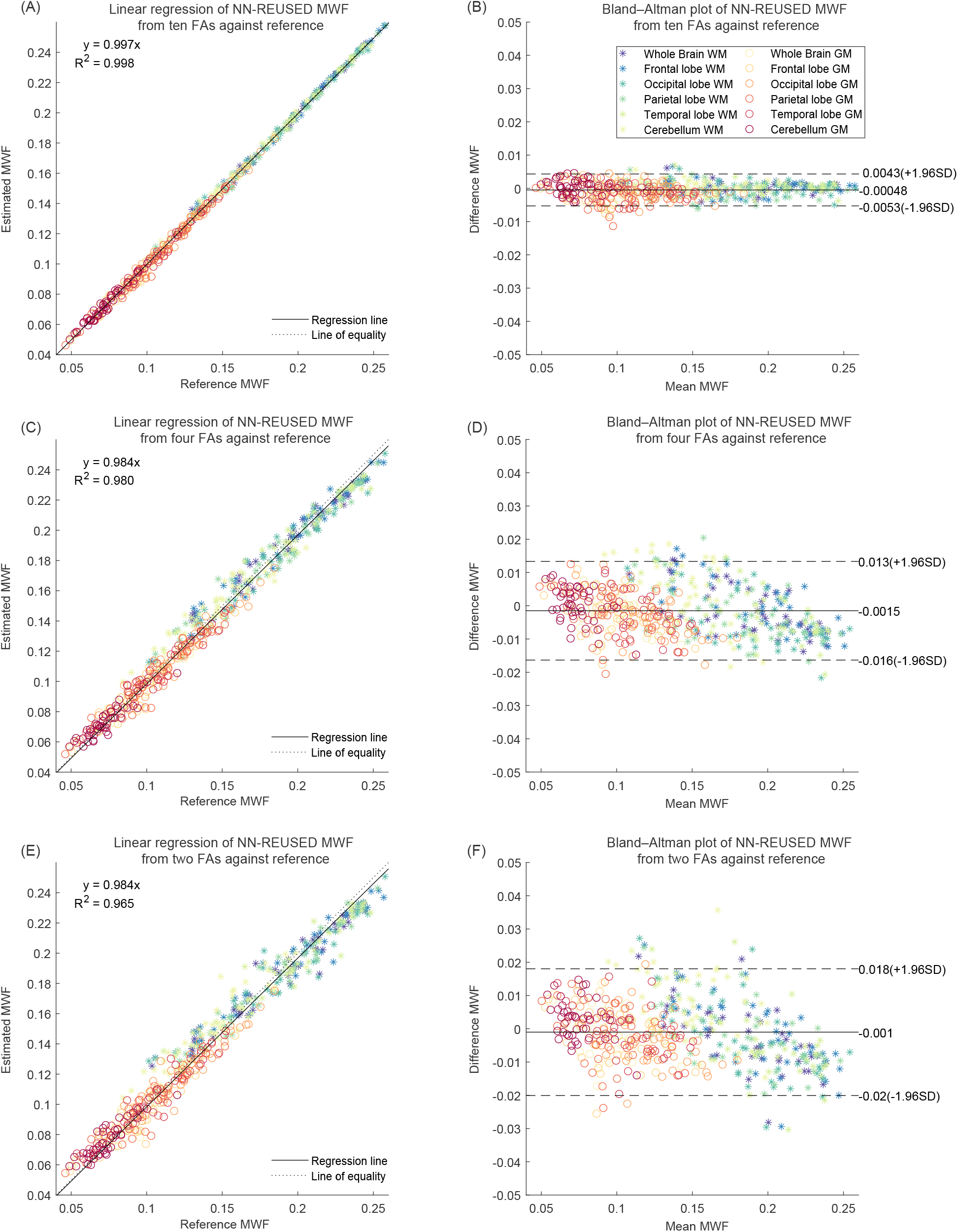
(A) Linear correlation plots between derived regional MWF values using REUSED and the reference method calculated across 12 WM and GM ROIs and across all testing datasets. (B) Corresponding Bland-Altman plots. Results are shown for the fully-sampled datasets (upper row), the under-sampled datasets (middle row), and the extremely under-sampled datasets (lower row).

**FIG. 6.**
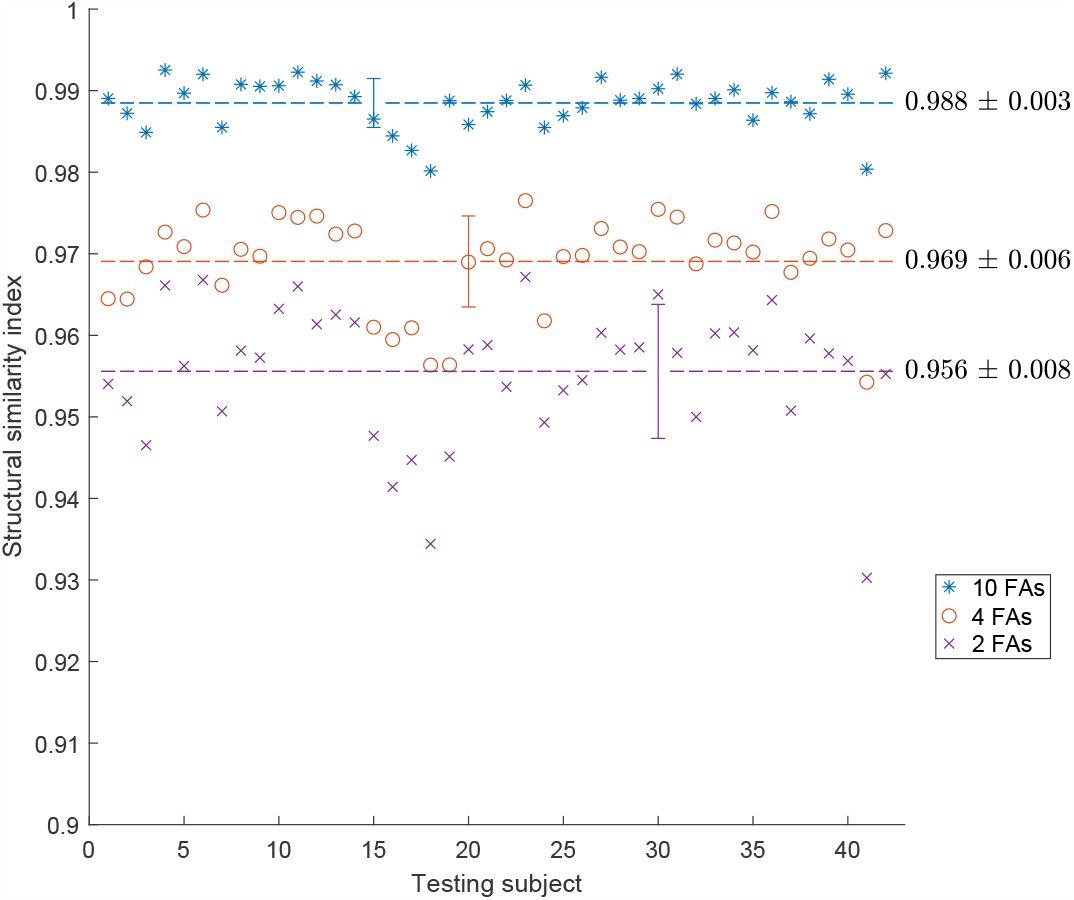
The structure similarity index measure (SSIM) calculated between derived MWF map using NN-REUSED and the reference MWF map for each testing subject. Mean and standard deviation of SSIMs calculated across all testing subjects are labeled using the dashed lines and error bars, respectively, for the fully-sampled (blue), under-sampled (orange), and extremely under-sampled (purple) datasets.

## IV. RESULTS

### A. NN-REUSED derived MWF maps from the fully-sampled datasets

Fig.2 shows examples of MWF maps derived using BMC-mcDESPOT or NN-REUSED from the fully-sampled datasets of an AD patient and a healthy subject. Visual inspection indicates excellent agreement between the MWF maps calculated using both approaches for both subjects. Interestingly, the patterns of demyelination in the AD subject are recovered with excellent fidelity using NN-REUSED. This further highlights the accuracy and utility of NN-REUSED since the training was conducted exclusively on datasets acquired from cognitively normal participants. Further, the very low AE values illustrate the nearly perfect regional determination of MWF values using NN-REUSED. Corresponding acquisition time (AT) and processing time (PT) for each method are also provided in the figure to demonstrate the substantial reduction in the PT using NN-REUSED as compared to BMC-mcDESPOT.

### B. NN-REUSED derived MWF maps from the under-sampled or extremely under-sampled datasets

Fig.3 shows examples of MWF maps derived using the reference method (i.e., BMC-mcDESPOT from the fully-sampled datasets), NN-REUSED or BMC-mcDESPOT from under-sampled datasets of an AD patient and a healthy subject. It is readily seen that while BMC-mcDESPOT fails to produce comparable MWF maps for under-sampled datasets, NN-REUSED derived MWF maps were visually similar to those derived using the reference method leading to great reductions in the AT and PT of, respectively, 10 min and 30 s, in comparison to the reference methods which requires an AT of 21 min and a PT of several hours. The high performance of REUSED is further highlighted by the low absolute error values in most brain regions. More interestingly, NN-REUSED performs nearly identically for extremely under-sampled datasets with marginal error introduced and a further reduction in AT Fig.4. In comparison, the BMC-mcDESPOT method could not produce meaningful MWF maps from extremely under-sampled datasets.

### C. Regional analysis of derived MWF values using NN-REUSED

The quantitative analysis of the NN-REUSED derived MWF results were conducted in 12 ROIs for both WM and GM and across all testing datasets. Linear regression plots and Bland-Altman plots of derived regional MWF values are shown in Fig.5. Here again, it is readily seen that derived MWF using REUSED from the fully-sampled, under-sampled or extremely under-sampled datasets exhibit very strong correlations with those derived using the references method, with correlation coefficients ranging from 0.97 to 0.99. Although the error in derived MWF values using REUSED increases with increasing the under-sampling of the datasets, these deviations remain significantly low, as clearly shown in the corresponding Bland-Altman plots. This analysis further demonstrates that NN-REUSED allows substantial reductions in both AT and PT with a minimal trade-off in estimation errors.

### D. Structure similarity of derived MWF maps using NN-REUSED

Providing a different metric than regional analysis, the perceived similarity of NN-REUSED MWF maps to the reference is examined using the structural similarity index (SSIM). Fig.6 displays the SSIM for each of the 42 testing subjects. The random distribution of the SSIM around the mean value across all testing datasets reflects the unbiased estimation of MWF maps using the NN-REUSED approach. Although the SSIM values decrease with increasing the under-sampling of the data, the values remain remarkably high with values of 0.988 *±* 0.003, 0.969 *±* 0.006, 0.956 *±* 0.008 for the fully-sampled, under-sampled, and extremely under-sampled datasets.

## V. DISCUSSION

In this work, we introduced a new NN model, REUSED, for the estimation of MWF, a surrogate of myelin content, from steady-state images, namely, SPGR and bSSFP, as incorporated in the BMC-mcDESPOT method^21^. Our results showed that NN-REUSED provides high accuracy and precision in derived MWF values. Indeed, the calculated MWF maps and regional values using NN-REUSED were virtually identical to those derived using the reference BMC-mcDESPOT approach. Importantly, NN-REUSED permitted estimation of MWF from undersampled and extremely under-sampled SPGR and bSSFP datasets, leading to a remarkable reduction in the total acquisition times with whole-brain high-resolution MWF map could now be generated within 7 min using NN-REUSED instead of 21 min as required by the reference method, BMC-mcDESPOT. Visual assessment of derived maps and quantitative analyses demonstrated the robustness of NN-REUSED to generate MWF values with high fidelity, accuracy and precision. In contrast, estimating MWF using the reference BMC-mcDESPOT approach with such limited datasets is not reliable, given the underdetermined nature of the fitting problem in this particular case. A higher number of SPGR and bSSFP images would be required for the large number of the unknown parameters to be jointly estimated in the BMC-mcDESPOT signal model^21^. We note that the total acquisition time could be further reduced using other methods for *B*_1_^+^ mapping such as steady-state double angle method^45^ or Bloch-Siegert shift^46^. These techniques are able to generate whole-brain *B*_1_^+^ maps within ∼1 min in contrast to ∼4 min using DAM; this will represent an additional substantial improvement of the temporal resolution for whole-brain high-resolution MWF mapping using NN-REUSED within 4 min. Another critical advantage of NN-REUSED is the computational speed. In fact, a whole-brain MWF map can be generated within a few seconds in contrast to ∼30 hours required for the BMC-mcDESPOT or the original mcDESPOT approaches. This drastic improvement in processing speed has several applications, including facilitating near real-time evaluation of the results as well as processing large datasets within a very short period of time without the need for expensive computational power.

Interestingly, NN-REUSED was able to generate virtually similar MWF maps, from either the fully-sampled, the under-sampled or the extremely under-sampled datasets, to those obtained using the reference method from the AD patient. We note that NN-REUSED was trained solely on datasets obtained from the brains of cognitively unimpaired participants. This further demonstrates the robustness and the applicability of this approach to a wide range of clinical investigations, including studies of the etiology and sequelae of myelin breakdown in neurodegeneration^17,47–49^. We conjecture that training these NN models on datasets that include patients suffering from demyelinating neuropathologies would further increase the accuracy and precision of NN-REUSED for MWF determination. Unfortunately, the lack of such datasets from our study cohort refrained us from conducting further analysis. However, this represents one of the future directions of this work.

Beyond the robustness and the substantial reduction in both the acquisition and computational times, NN-REUSED will allow conducting of complementary analyses of previous investigations where only a limited number of SPGR and bSSFP images were acquired. Indeed, the SPGR and bSSFP images acquired at a limited number of FAs can be used to generate MWF maps and, therefore, to derive further insights into the myelination patterns of the previously studied condition. Further, in numerous precedent mcDESPOT or BMC-mcDESPOT investigations^12,21,25,26,50,51^, several datasets were excluded from the underlying analyses due to motion artifacts in certain SPGR and bSSFP images. Using NN-REUSED, these datasets could potentially be incorporated to derive corresponding MWF maps from the remaining SPGR and bSSFP images that are free of motion artifacts. The incorporation of such datasets is likely to improve the statistical power of those underlying analyses.

We have also successfully implemented and tested various other NN models, including conventional CNN, U-Net^52^ and cGAN^53,54^, but faced several limitations. One of these major drawbacks was the prolonged time for the training; this is due to the much larger number of calculations needed for the convolution operations. In fact, several days were required for the convergence of CNN, cGAN, and U-Net in order to produce reasonable results. This is in contrast to only a few hours for the fully connected NN models. Further, and more importantly, derived MWF maps using the CNN and U-Net models exhibited substantial blurring, especially around tissue edges and small structures (data not shown). Therefore, despite its simplicity, the fully connected residual NN model provided superior results; this is likely due to a large number of available training features (∼78 million).

This work comes with limitations. Despite the accuracy of the NN-REUSED approach introduced here to robustly estimate MWF from data of normal or AD subjects, further testing is required using a larger dataset that includes various degrees of demyelination or neuropathologies. Furthermore, as with all machine learning-based algorithms, a training dataset is initially required; this can represent an actual challenge, especially when the expertise or infrastructure is lacking to construct this training dataset. However, for the case of the BMC-mcDESPOT analysis, our training dataset and algorithms are available and readily applicable to other studies. Finally, NN-REUSED was tested on steady-state imaging only. Further analyses are required to test this proof-of-principle on other imaging modalities of MWF, including those based on multi-spin echo or multi-gradient echo MR sequences^37,55,56^.

## Data Availability

All data produced in the present study are available upon reasonable request to the authors

## ACKNOWLEDGMENTS

This work was supported by the Intramural Research Program of the National Institute on Aging of the National Institutes of Health. This work utilized the computational resources of the NIH HPC Biowulf cluster (http://hpc.nih.gov).

